# Incorporating Real-World Clinico-Genomic Insights to Inform Diversity Enrollment Targets in Oncology Trials

**DOI:** 10.1101/2024.04.17.24305991

**Authors:** Francisco M. De La Vega, Yannick Pouliot, Brooke Rhead

## Abstract

The passage of the US Food and Drug Administration (FDA) Omnibus Reform Act of 2022 underscores a national commitment to enhancing diversity in clinical trials. This commitment recognizes not only the ethical imperative of inclusivity but also the practical necessity to ensure the safety and efficacy of medications across all demographic groups. Particularly for Phase 3 and pivotal clinical trials, the FDA has issued draft guidance that recommends sponsors to develop diversity plans with race and ethnicity (R/E) enrollment targets informed by the epidemiological landscape of the disease in the therapy’s target population. For biomarker-driven oncology trials, real-world data (RWD), especially when enriched with multimodal clinico-genomic information, holds immense promise for informing these R/E enrollment goals.

However, leveraging RWD comes with hurdles, including the overrepresentation of insured patients, significant non-random missingness in R/E data, and disparities between R/E distributions in RWD and disease incidence databases—often attributed to healthcare access and socioeconomic disparities. Here, we propose a robust methodology to harness clinico-genomic RWD, addressing these challenges through strategies that include accurate R/E imputation and incidence adjustment factors. Our approach then utilizes clinical data and biomarker prevalence in RWD to derive a data-driven R/E distribution for clinical trial enrollment targets.

Through a case study on a hypothetical biomarker-driven clinical trial targeting prostate adenocarcinoma and leveraging a cohort from the Tempus clinco-genomic database, we demonstrate the application of our methodology. This example illustrates the potential of RWD to offer enrollment target scenarios, grounded in disease epidemiology and empirical R/E distributions adjusted for biomarker prevalence. Such data-driven targets are pivotal for the development of informed and equitable diversity plans in oncology clinical trials, paving the way for more representative and generalizable research outcomes.

## Introduction

The recognition of racial and ethnic underrepresentation in clinical trials by regulatory agencies highlights a critical gap in our healthcare system, directly linked to persistent health inequities.^1^ Recent assessments have shown notably lower participation rates among Black and Hispanic/Latino patients compared to their White counterparts, across multiple cancer types.^2^ This discrepancy is compounded by the frequent absence of disaggregated race and ethnicity (R/E) data in clinical trial reports^3^ obscuring the ability to monitor and address these disparities effectively. This lack of diversity not only challenges the generalizability of trial outcomes but also perpetuates existing health inequities by limiting the ability to deliver treatments that benefit diverse patient populations.^4^

Moreover, the introduction of the Food and Drug Administration (FDA) Omnibus Reform Act of 2022 (FDORA) signifies a legislative push towards diversifying clinical trial participation^1^. As mandated by this legislation, in 2022 the FDA has issued draft guidance recommending the submission of diversity action plans for Phase 3 and pivotal trials, covering drugs, biological products, or devices, highlighting the FDA’s commitment to modernizing clinical trial diversity.^5^ These plans are expected to encompass not only R/E enrollment goals but also strategies encompassing patient-directed measures, community engagement, workforce-directed measures, and trial design modifications aimed at overcoming barriers to participation, including geographic and socioeconomic disparities.^5,6^

Given the shift towards biomarker-driven drug development in oncology,^7–9^ the utilization of real-world data (RWD), particularly clinico-genomic databases, presents a promising avenue for informing R/E enrollment goals.^10^ However, a number of issues create challenges for effectively leveraging RWD for this purpose, such as the overrepresentation of insured and White patients,^11^ significant non-random missingness in R/E data,^12–16^ and inconsistencies between RWD and disease incidence databases.^17^ These hurdles often reflect broader issues of healthcare access and socioeconomic disparities.^18^ Additionally, R/E data in RWD may originate from various sources, not exclusively self-identified R/E, but also includes the assignment of R/E classifications on patients by third parties.^19^ Furthermore, R/E missingness in RWD can be attributed to issues in data collection and transmission rather than simply patient abstention.^12,20,21^

To navigate these obstacles, we propose a robust methodology leveraging clinico-genomic RWD, incorporating an accurate R/E imputation method^13^ and incidence adjustment factors to derive data-driven R/E distributions for use in the development of clinical trial enrollment targets. Our methodology exemplifies how such an approach can be applied through a case study on a hypothetical biomarker-driven clinical trial targeting prostate adenocarcinoma, leveraging data from the Tempus clinico-genomic database. This case study highlights the potential of RWD to inform enrollment target scenarios that are not only based on the epidemiology of the disease but also adjusted for empirical R/E distributions and biomarker prevalence, thereby supporting the development of informed and equitable diversity plans in oncology clinical trials.

## Methods

### Patient cohorts

We obtained data from the de-identified Tempus clinico-genomic database, which includes genomic and clinical data from cancer patients that underwent tumor profiling using Tempus’ xT assay as part of their healthcare.^22^ Selection criteria included tumor profiling with the Tempus xT assay (v2-v4) from 2018 to 2022, selecting the test with the first collection date in case of multiplicity. For our analyses of disparities between expected vs observed R/E distribution for the top ten most frequent cancers in the Tempus database, we selected a cohort of 41,856 deidentified patient records. For our mock clinical trial case study, a cohort of 4,328 patients diagnosed with prostate adenocarcinoma with sequencing of prostate gland tumor tissue was selected. Demographic information included: patient age at date of specimen collection, age at diagnosis, gender, and stated (i.e., either self-reported or observed) R/E. Clinical information included: histology, PSA measurements, stage, grade, total Gleason score (raw and aggregated), and castration resistant status derived from clinical records (cf. Supplementary Table 1).

### Definition of race and ethnicity categories

The R/E classifications in EHR, cancer statistics, and RWD sources in the US, follow the 1997 federal guidelines from the US Office of Management and Budget. ^25^ These guidelines encompass two self-reported categories: a) Race (options include American Indian or Alaska Native, Asian, Black or African American, Native Hawaiian or Other Pacific Islander, and White); and b) Ethnicity (options include Hispanic or Latino and Not Hispanic or Latino).^26^ To address analytical challenges due to overlapping race and ethnicity categories, our study opts to consolidate responses into mutually exclusive categories:^26^ Hispanic or Latino, non-Hispanic (NH) Asian, NH Black, and NH White, noting that other racial groups currently lack sufficient representation in our data to support robust model development.^26^ The federal R/E category standards have recently been updated^27^ although their implementation is not expected until 2029.^27^ Self-identified R/E (SIRE) is considered the gold standard R/E data. However, RWD may include R/E data from third parties, healthcare providers, or others who either assign categories based on physical characteristics,^19^ or omit this data, either unintentionally due to error, or intentionally to prevent discrimination.^28,29^ Therefore, we refer to R/E data in RWD as “stated” R/E rather than SIRE.^13^

### Imputation of race and ethnicity categories

To overcome missingness of stated race and ethnicity in our real-world data, we performed imputation of mutually exclusive R/E categories with a previously reported method.^13^ The assessment of the sensitivity and specificity of the heuristic version of method used here,^13^ demonstrated high accuracy with data from the Tempus database (correct rate of 96% and weighted error of 0.9% ^13^), performing much better than other commonly used methods, such as the Medicare Bayesian Improved Surname Geocoding^31^ (MBISG; reported correct rate of 78% and weighed error of 8.9%^32^), and with low no-call rate (∼3%).^13^

### Disparity between expected vs observed R/E distribution in the Tempus database

We calculated the difference between the expected and observed distribution of R/E categories by first determining the expected distribution from newly diagnosed cancer cases data reported in the USCS Data Visualizations Tool^33^ data tables for the years 2015-2019 (release date November 2022; https://www.cdc.gov/cancer/uscs/USCS-1999-2019-ASCII.zip). We conducted our analysis at the state level to overcome frequent R/E data suppression at the county level in the USCS data, and further limiting to states where none of the R/E categories we analyzed were censored and to states within the service area of Tempus. “Unknown” race and NH American Indian and Alaska Native categories in the USCS data were excluded from our analysis.

For each state we calculated *E_c,r,s_*, the expected proportions of patients with cancer *c* for race-ethnicity *r* in state *s* as: counts per race-ethnicity/total patients for cancer *c* in state *s* in the USCS data. Similarly, we calculated *O_c,r,s_*, the observed Tempus proportions of patients of cancer *s* for race-ethnicity *r* in state *s* as Tempus counts per cancer per race-ethnicity/Tempus total patients for cancer *c* in state *s*. We then calculated the disparity fraction^34^ (*DF_c,r,s_*), the difference between the expected proportion and the observed proportion in the Tempus data for each cancer, race-ethnicity, and state:

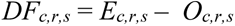

If *DF_c,r,s_* > 0, group *r* is under-represented, whereas if *DF_c,r,s_* < 0, group *r* is over-represented.

We further calculated overall *DF*s for each cancer, weighting each state-level *DF* by the proportion of cases in the USCS data from that state and the proportion of Tempus data obtained from that state as follows. First, we calculated the USCS weight for cancer *c* in state *s*: *W_c,s_*= USCS incidence count of cancer *c* in state *s* / USCS total incidence counts for cancer *c*. Next, we calculated the Tempus sampling rate of cancer *c* in state *s*: *SR_c,s_*= counts of Tempus patients for cancer *c* in state *s* / total number of cancer *c* patients in selected Tempus cohort. We computed the adjusted weight for state *s* in cancer *c* as: *AW_c,s_= W_c,s_*SR_c,s_*.

We then calculated the weighted disparity fraction for each cancer as:

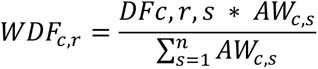

### Statistical significance of differences between expected and observed R/E distributions

We performed a binomial proportion test to determine whether observed differences were statistically different to expected, per cancer, per R/E, per state. Then we aggregated *p*-values from the binomial proportion tests across states, per cancer, per R/E, by weighting each state-level *p*-value as before by *AW_c,s_*, using Stouffer’s Z-score method. To make *p*-values equivalent to 2-tailed *p*-values, we select the minimum of the “greater” and “less” combined *p*-value for each R/E, we multiplied each *p*-value by 2, and if any of the resulting *p*-values was greater than 1, we replaced by 1. We adjusted the aggregated *p*-values for multiple testing with the Benjamini-Hochberg procedure for controlling the false discovery rate on the combined 2-tailed *p*-values. The number of tests was the number of cancers times the number of R/E categories (Supplementary Table 2).

### Adjustment factors for enrollment targets

Let *O*′*_c,r_* represent the new distribution of R/E categories for the cancer type under study *c* for each R/E category *r* after applying the inclusion/exclusion criteria (I/E). To adjust these new proportions (*O*′*_c,r_*) for the initial disparity, we use the weighted disparity fraction (*WDF_c,r_*). However, since *O*′*_c,r_* may represent the distribution of a disease subtype, the adjustment needs to be made carefully to acknowledge the reasons for these criteria while also addressing the initial disparity. We proportionally adjust *O*′*_c,r_* by a factor derived from *WDF_c,r_* ensuring that the adjustments do not counteract the inclusion criteria’s purpose. This adjustment aims to balance the need to respect the impact of the inclusion criteria with the goal of mitigating initial disparities. Hence, we define *A_c,r_*, the adjusted final proportion, as follows:

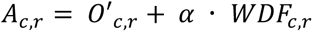

where *α* is a scaling factor that determines how strongly to adjust *O*′*_c,r_* based on the initial disparity fraction. The scaling factor *α* is normally set at 1, nonetheless it is abrbitrary and depends on the weight given to correcting for disparities versus adhering to the new proportions dictated by the inclusion criteria. Once *A_c,r_* is obtained for each R/E category, we calculate the total of the adjusted proportions and normalize them to ensure they sum to 100%, *Ã_c,r_* – this can be done simply by rounding. Finally, one can multiply *Ã_c,r_* with the total trial enrollment target for each R/E to obtain initial diversity enrollment targets.

## Results

### Assessment of disparities in R/E proportions between national cancer incidence data and RWD

We reasoned that differences in R/E distribution between epidemiological incidence databases and RWD can inform about specific over-or under-representations of R/E categories in RWD sources, enabling the development of correction factors. Calculating the expected distribution based on incidence data is complex, as incidence varies significantly by county due to environmental and socioeconomic factors,^33,35^ and suppression/censoring of some R/E categories in some counties due to privacy reasons, such that a given RWD may not represent all US counties or states equally. Expected R/E distributions can be calculated using cancer incidence data from databases such as the Surveillance, Epidemiology, and End Results (SEER) program,^23^ or the CDC’s US Cancer Statistics (USCS) database.^24^ The latter offers a more comprehensive overview by including data from most US states and incorporating SEER and the National Program of Cancer Registries (NPCR) data.^24^ Since cancer incidence can significantly vary between counties,^35^ comparing RWD with nationwide incidence data may not always be appropriate. Further, for clinico-genomic databases derived from clinical genomic testing, it is crucial to consider the patient catchment area and apply weights to adjust for sampling variances across counties.^36^ In the Methods section, we describe a procedure to calculate such expectations for different cancer types reported in the USCS and determine whether there are statistically significant differences between the expected and observed distributions.

By comparing expectations with observed R/E distributions, we calculate a weighted disparity fraction (*WDF*) to reveal representation biases by R/E in specific cancer types.^34^ Figure 1 illustrates the R/E disparity fractions for 10 major cancer types showcasing the varied over-and under-representations across cancers. Some disparities are statistically significant, and while the disparities are small (less than 8 percent points overall), unexpected patterns can 1be observed. For example, in our pan-cancer cohort, we observed an over-representation of NH Asian patients in breast cancer cases, and Hispanic/Latino patients in lung cancer and melanoma cases. Conversely, under-representation was noted among NH Black patients with gastroesophageal cancers, Hispanic/Latino patients with prostate cancer, and NH White patients with breast, colorectal, melanoma, and pancreatic cancers. The latter may seem counterintuitive; however, considering that tumor profiling is not yet common in first-line therapy for many cancers,^37^ and that most patients undergoing tumor profiling are in stages 3 or 4 (Supplementary Table 1), we hypothesize a depletion of regularly screened patients diagnosed in earlier stages when the disease is more curable. For instance, in melanoma cases, detecting tumors is easier in patients with lighter skin and the disease is more easily cured in early stages.^38^

**Figure 1.**
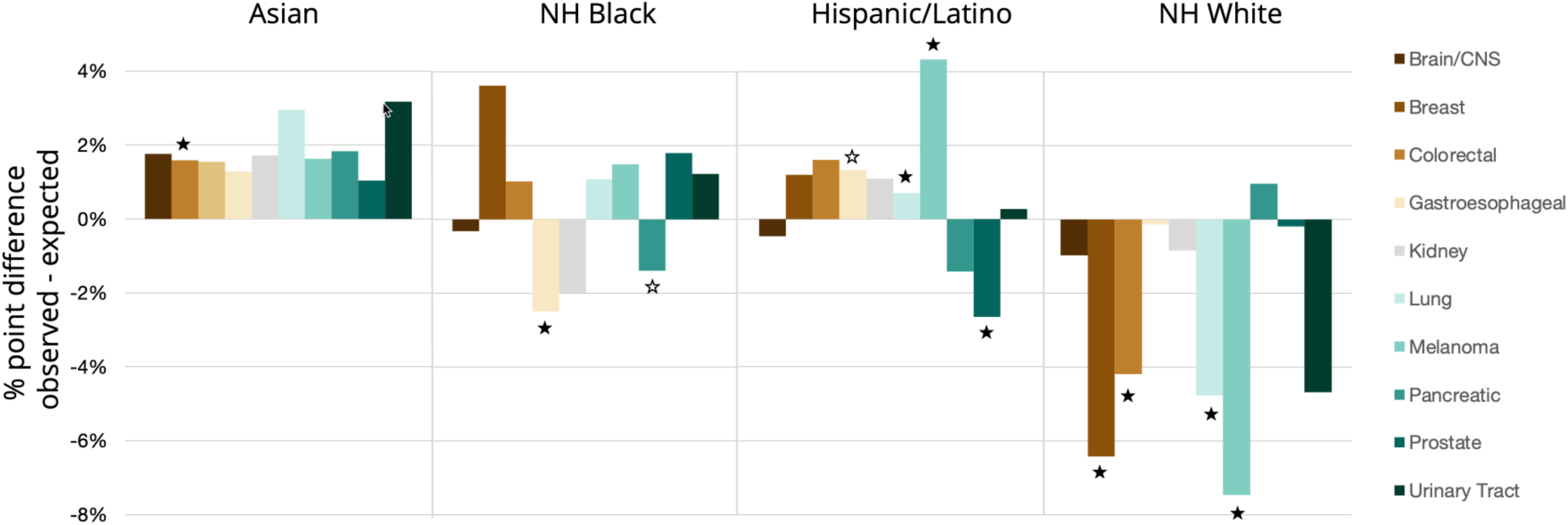
Racial/ethnic disparities in the distribution of patients sequenced per cancer type with respect to United States Cancer Statistics (USCS) database of cancer incidence. We looked for differences between the observed distribution of racial/ethnic categories per cancer type in our cohort, and the expectation based on cancer incidence rates from the USCS database between 2015-2019 at the state level, rolled up as a weighted average adjusted by our sampling rate (number of patients in our cohort from each state; cf. Methods and Supplementary Table 3). Sample sizes: Asian = 2,733; NH Black = 7,168; Hispanic/Latino=5,252; and NH White = 44,464. We performed one proportion Z-test to assess the differences between observed and expected proportions of race/ethnicity categories at the state level. We aggregated p-values across states using Stouffer’s Z-score method. A star indicates statistically different differences from expectation (p<0.05) - open star nominal value, black start after multiple testing adjustment (cf. Supplementary Table 2).

### A workflow to establish R/E enrollment targets in oncology trials from clinico-genomic RWD

With the above considerations in mind, we propose a workflow to establish data-driven R/E enrollment targets for oncology clinical trials, leveraging data from de-identified clinico-genomic databases. The workflow, detailed in Box 1 and illustrated in Supplementary Figure 1, is composed of three major steps: I) Assess disparities between expected and observed R/E in the chosen RWD source; II) Modeling the impact of I/E criteria in R/E distribution, including biomarkers, to finalize the selection of I/E criteria; and III) Compute enrollment targets, rectifying differences between expected and observed R/E distributions in RWD.

#### Box 1.

##### A workflow to establish R/E enrollment targets from RWD

Step I – Assess disparities between expected and observed RWD R/E distributions.

1. Obtain expected distributions from incidence databases such as USCS.
2. Evaluate the disparity between expected (based on incidence data) and observed R/E distributions for the specific RWD cohort used in modeling.
3. Assess whether these differences are statistically significant. These assessments can be done using subsets of the cohort with complete stated R/E data (complete case analysis), or the entire cohort by using imputed R/E data.
4. If significant differences are observed, derive correction factors to be applied later in the workflow. Given that R/E missingness significantly reduces the sample size available for the analysis and can be biased, imputed R/E data is usually more reliable for this step.

Step II – Modeling of I/E criteria and its impact on R/E distributions.

1. Stratify the selected cohort by those clinical inclusion/exclusion (I/E) criteria that are available in the RWD database to generate post-I/E R/E distributions. Apply I/E criteria individually to determine which criteria have the most significant impact on R/E distributions.
2. If distributions are computed for both stated and imputed R/E data, compare them and assess whether there are differences. If differences observed between these are significant, a bias in missingness may be present, and imputed R/E may be more reliable. Caution is needed with stated R/E if, after applying I/E criteria, the sample size of underserved populations is too small. These considerations allow sponsors to decide whether to rely on stated R/E, or instead proceed with imputed R/E.
3. For biomarker-driven clinical trials, assess the impact of biomarker presence on R/E distributions. This analysis will aid in reassessing I/E criteria and, if possible, in avoiding criteria or clinical thresholds that unnecessarily reduce participation from underserved minorities. The analysis above can be applied to both stated and imputed R/E.

Step III – Compute R/E enrollment targets.

1. Once I/E criteria have been finalized, combine these criteria to compute a final R/E distribution based on RWD.
2. If Step I resulted in significant differences between expected and observed R/E distributions at the cohort level, the correction factors developed in Step 1 may be applied to obtain a final R/E distribution to derive enrollment targets. Scaling factors can be used to balance adjustment vs. other goals.

### A case study to demonstrate our workflow: Hypothetical prostate cancer trial design

To illustrate the application of our workflow with a practical example, we will navigate through the steps of the process for a hypothetical interventional trial targeting prostate cancer treatment. Prostate cancer, a disease characterized by significant racial and ethnic disparities, is known to disproportionately affect Black men in the United States and globally^39^. Despite the population-level incidence rate being nearly 1.8 times higher in Black men compared to White men^40^, clinical trials often fail to accurately represent these disparities among R/E subgroups^39^. This underrepresentation highlights the importance of prostate cancer as an exemplary case for demonstrating how our methodology can improve the participation of underrepresented minorities in oncology clinical trials by establishing data-driven enrollment targets.

Our case study involves a hypothetical interventional trial aiming to enroll 500 men with stage 3 or 4, ETS-positive prostate adenocarcinoma (PRAD). The ETS gene family, comprising 28 transcription factors, frequently shows aberrant expression in prostate cancer, with *ERG* being the most commonly affected^41^ The overexpression of *ERG*, mainly due to structural rearrangements of the transmembrane protease serine 2 (*TMPRSS2*) with *ERG* (with *ETV1* and *ETV4* being less common), is a hallmark in over 30% of prostate cancer cases.^42^ Our scenario relies on the notion of repurposing of drugs that may be effective in ETS-positive cancers.^43^ Specifically, our hypothetical scenario considers a repurposed drug therapy being tested in a trial for patients with prostate adenocarcinoma confirmed to have TMPRSS2:ERG gene fusions. As tumor profiling becomes increasingly common in the cancer treatment journey,^44^ the feasibility of biomarker-driven clinical trials targeting specific molecular alterations is on the rise. Real-world, clinico-genomic databases offer multimodal clinical and genomic data useful in modeling biomarker-driven clinical trials. As we demonstrate below, they also provide insight into setting R/E enrollment targets and developing diversity plans for such trials.

### Set-up: RWD cohort and eligibility criteria

The RWD for this analysis was obtained from the Tempus de-identified clinico-genomic database.^22,45^ We selected a cohort of 4,328 PRAD patients that underwent tumor genomic profiling of prostate tumor tissue with the Tempus xT assay.^22^ Supplementary Table 1 shows the cohort patient characteristics by imputed race and ethnicity. We propose a list of tentative I/E criteria for participation in our hypothetical trial in Box 2. This list is a basic set of criteria that can be readily explored in RWD, sufficient to exemplify our process.

#### Box 2.

##### Candidate Eligibility Criteria

Inclusion Criteria:

- Histological proof of adenocarcinoma of the prostate.
- Detectable PSA of at least 2µg/ml.
- Prostate biopsy histology grade total Gleason ≥ 6.
- Stage 3 or 4 disease.
- Presence of TMPRSS2:ERG gene fusions assessed centrally by a gene mutation biomarker panel.

Exclusion Criteria:

- Pathological findings consistent with small cell carcinoma of the prostate.
- Known castration-resistant disease.

### Expected vs observed R/E distribution differences for the prostate cancer cohort (Step I)

As described in Step 1 of our workflow and detailed further in the Methods section, we utilized the USCS database to obtain the expected distribution of R/E for prostate cancer patients. We then calculated the difference in proportions between the expected R/E distributions and those observed in our cohort, (the weighted disparity fraction, *WDF*). Table 1 presents these results.

**Table 1.**
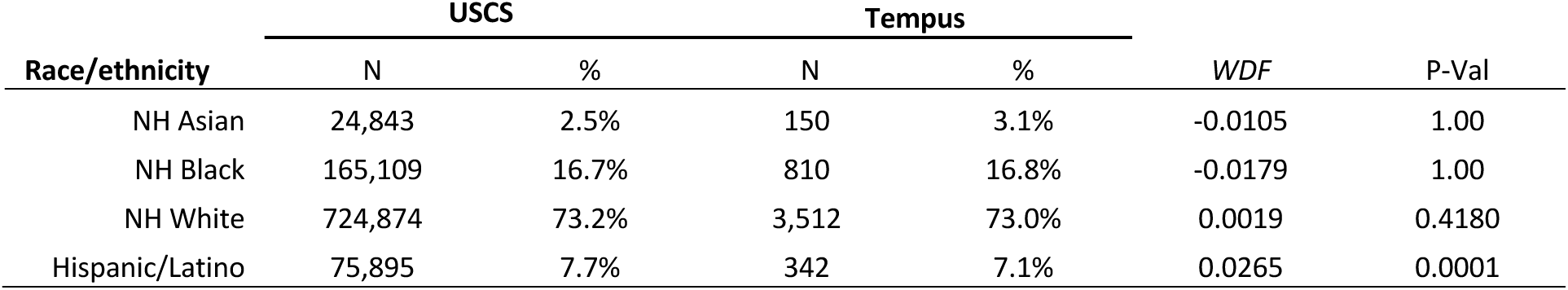
Distributions of race and ethnicity categories in USCS and Tempus and the weighted disparity fractions *(WDFs*) for each category are presented. Note that in over-represented categories, *WDF*s are negative, whereas in under-represented categories, these values are positive. Statistical significance of *WDF*s were evaluated using a binomial proportions test (refer to Methods and Supplementary Table 2 for details).

Table 1 reveals that, following our normalization procedure, there are no significant differences between the expected and observed proportions for NH Asian, NH Black and NH White patients. However, there is a small statistically significant underrepresentation of Hispanic/Latino patients (2.65%) in our Tempus prostate cancer cohort. Since there is at least one significant difference, these fractions will be used in Step III as correction factors to adjust final R/E distributions after applying I/E factors to set adjusted enrollment targets.

### Assess impact of I/E criteria on R/E distributions (Step II)

The next step involves assessing the impact of I/E criteria on the R/E distributions found in RWD. Table 2 presents the stated and imputed R/E distributions for different strata of our PRAD cohort. The first column displays the R/E distribution for patients with histologically confirmed PRAD. The subsequent columns reveal the R/E distributions when applying individually various inclusion criteria: stage 3 and 4, total Gleason score ≥6, and patients with at least one PSA measurement of ≥ 2µg/ml.

**Table 2.**
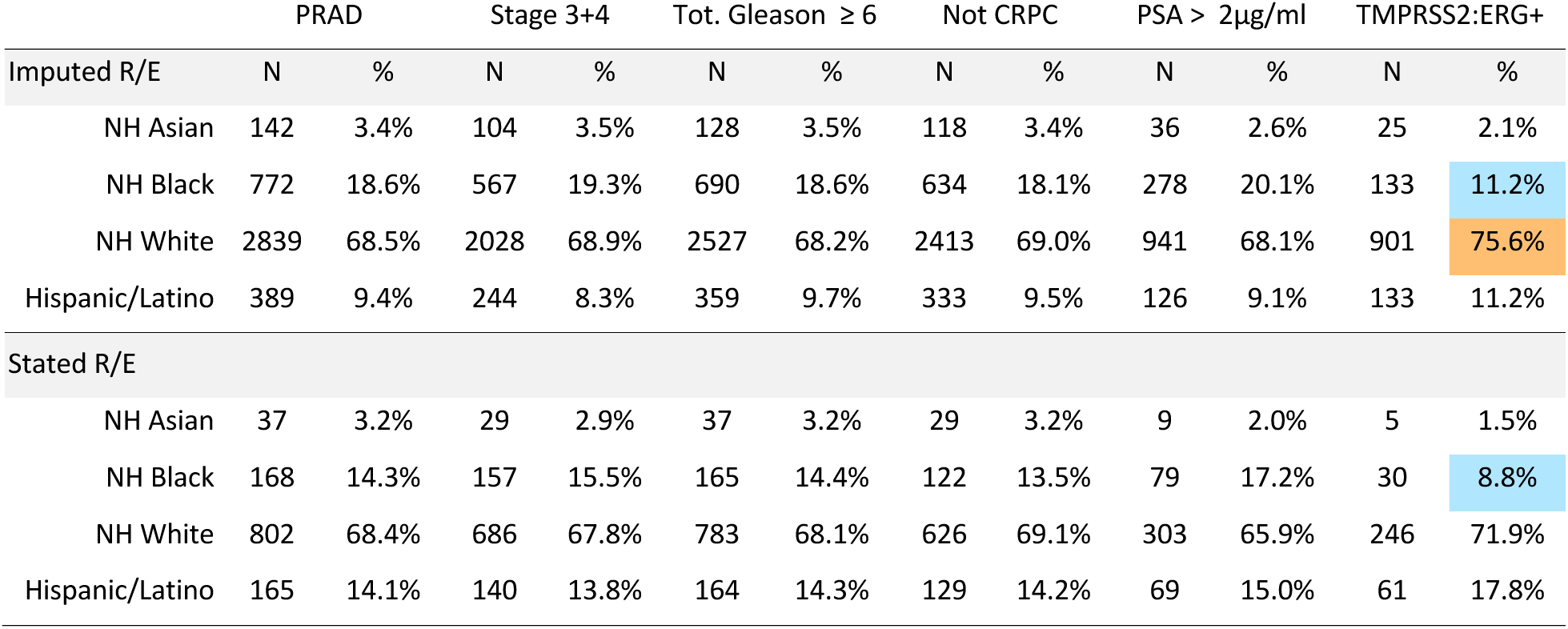
Distribution of race and ethnicity categories (R/E) across different inclusion criteria strata. Cells color highlight changes of five percentage points or more over (orange) or below (blue) the R/E distribution in the initial PRAD cohort. The top section presents data for imputed R/E categories, whereas the bottom section presents data for patients where stated R/E was available.

To illustrate the benefits of R/E imputation, Table 2 includes analyses for both imputed and stated R/E. Due to data missingness and the necessity of having available stated race and ethnicity metadata to produce mutually exclusive R/E categories, the counts of patients for each I/E stratum is significantly decreased when relying on stated R/E. This reduced sample size, coupled with the potential for bias in R/E missingness, diminishes confidence in the proportions derived solely from stated R/E.

Table 2 illustrates the impact of different inclusion criteria on the R/E distribution based on either stated or imputed data. We observe an over-representation of NH White patients with TMPRSS2:ERG gene fusions and under-representation of NH Black patients, a disparity noted in the literature for this biomarker.^46^ Notably, there are differences in sample size between imputed and stated R/E data, with missingness in stated R/E leading to less reliable figures for some groups. Furthermore, the stated R/E distributions show less pronounced disparities compared to imputed data (e.g., the over-representation of NH White individuals in the TMPRSS2:ERG+ group is reduced by half), suggesting potential bias in missingness (given the reported accuracy of our imputation method).^13^

Another factor to consider is how each I/E criterion reduces the sample size available for analysis. Specifically, when requiring a PSA measurement of >= 2µg/ml, only 33% of the initial PRAD cohort meets this criterion, partly due to the 37% missingness in PSA measurements. The cause of this missingness is unknown, but we cannot rule out the possibility that it results from biases in healthcare access, or other socioeconomic factors associated with R/E, rather than inherent differences in the tumors of the patients in these categories. Therefore, we decided to eliminate this criterion in the final step to define R/E distributions for setting enrollment goals.

### Define R/E enrollment targets (Step III)

Once we have evaluated the impact of the different inclusion criteria on the R/E distribution, these can be examined to assess whether any of them create unnecessary disparities and can be eliminated. Some criteria are necessary for the therapy in question and will remain (e.g., TMPRSS2:ERG positive tumor). Once the I/E criteria are finalized, they can be combined to obtain a final distribution of R/E in RWD for the desired patient population (Table 3). This allows us to contrast this distribution with the initial PRAD cohort and with the expectations from the USCS incidence data as derived in Step I.

**Table 3.** Development of initial and adjusted R/E enrollment targets based on I/E criteria and the disparity fraction between the RWD and US Cancer Statistics (cf. Table 1). We present two scenarios, the first with the scaling factor α is set to 1, and the second section displays results when it is set to 0.5. Cell colors highlight changes of five percentage points or more over (orange) or below (blue) the R/E distribution in the PRAD cohort.

| | Tempus Prostate | Tempus PRAD | I/E $O'_{c,r}$ | Initial target | WDFA | $\alpha = 1$ | | $\alpha = 0.5$ | |
| --- | --- | --- | --- | --- | --- | --- | --- | --- | --- |
| | | | | | | Adjusted ($A_{c,r}$) | Adjusted target | Adjusted ($A_{c,r}$) | Adjusted target |
| Imputed R/E | % | % | % | N | % | % | N | % | N |
| NH Asian | 3.1% | 3.4% | 2.3% | 11 | -1.1% | 1.2% | 6 | 1.8% | 9 |
| NH Black | 16.8% | 18.4% | 10.9% | 54 | -1.8% | 9.1% | 46 | 10.0% | 50 |
| NH White | 73.0% | 68.9% | 76.3% | 381 | 0.2% | 76.5% | 382 | 76.4% | 382 |
| Hispanic/Latino | 7.1% | 9.3% | 10.5% | 53 | 2.7% | 13.2% | 66 | 11.8% | 59 |

Table 3 shows the workflow to define R/E enrollment goals. It shows the distribution of R/E across the selected Tempus prostate cohort (N=4,814), the PRAD subset of that cohort (N=4,196), and *O*′_!,#_ – the observed R/E distribution after applying the final I/E criteria (N=523): PRAD, stage 3 or 4, total Gleason ≥6, and TMPRSS2:ERG+ as inclusion criteria, and applying confirmed CRPC as exclusion criteria. We then adjust for the disparity between the overall prostate cancer cohort in our RWD and the expectation derived from the US Cancer Statistics data (cf. Table 1). Following the Methods, we apply *WDF* as a correction factor, multiplied by a scaling factor. This adjustment results in a revised R/E proportion distribution and enrollment targets, with increases in Hispanic/Latino patients and minimal changes in the other groups. We also show that the impact of changing the scaling factor from 1 to 0.5 is very small.

## Discussion

Real-world clinico genomic databases —which include multimodal genomic data (e.g., DNA alterations and gene expression values)^22,48^ linked with diverse clinical data from electronic health records (EHR) or abstracted clinical documents—offer numerous benefits for understanding the epidemiology of diseases in real patient populations and their distribution across R/E categories. Particularly in modern biomarker-driven oncology clinical trials, clinico-genomic databases can be used to understand the prevalence of molecular biomarkers with respect to R/E.^45,49^ With tumor genomic profiling becoming increasingly common in clinical cancer care and recommended in treatment guidelines,^44^ the magnitude of these data is expanding rapidly. Despite representation biases, because of its scale, these data provide significant statistical power for analyses across all major R/E categories including underserved minorities.^50^ However, RWD often exhibits inconsistencies and missingness, especially regarding R/E data, with incompleteness rates varying from 30-80% depending on the source.^12,51^ Some of this missingness is not random;^12–15^ underserved populations tend to provide self-identified R/E (SIRE) less frequently.^29^ This gap significantly impacts the ability to define enrollment goals that align solely with disease epidemiology and biomarker prevalence.

In this paper, we outline a robust methodology utilizing real-world data to establish diversity enrollment targets for oncology trials, specifically emphasizing the use of clinico-genomic databases in biomarker-driven studies.^45^ We present strategies to address RWD challenges, such as missing R/E data and healthcare access biases, aiming to create fairer diversity plans. Our example of a prostate cancer trial demonstrates the application of this methodology, highlighting the refinement of R/E enrollment targets through the evaluation of disparities and imputation techniques. This approach sheds light on how inclusion/exclusion criteria affect R/E distribution, particularly in the context of molecular biomarkers where racial biases in their distribution may stem from various poorly understood factors. Data-driven approaches for eliminating unnecessary I/E criteria have been proposed.^52^ Our research suggests that these methods could be expanded to consider the impact of I/E on the distribution of R/E, especially when data missingness in criteria is present and could be biased by R/E.^46^

An important aspect of our methodology is conducting a thorough assessment of the disparities between expected R/E distributions in epidemiology or disease incidence data and those observed in a RWD source. It’s crucial to acknowledge that cancer incidence can vary significantly by geography.^35^ This variation, influenced by factors such as genetic predispositions, lifestyle choices, socioeconomic status, access to healthcare, and environmental factors,^53^ necessitates performing assessments with as much geographical granularity as possible, considering the catchment area of patients contributing to the RWD. This approach allows us to derive a weighted disparity fraction, which is instrumental in adjusting for the over-or under-representation of specific R/E groups in the RWD. These disparities are the result of a complex interplay of factors, including access to care (particularly early-stage curative therapies) and insurance status.^11,38^

Another important feature of our method is the use of imputation to address the R/E missingness problem in RWD. Several methods exist to impute R/E from clinical administrative data, such as the widely used Bayesian Improved Surname and Geocoding method (BSIG).^31^ Machine learning methods that leverage EHR data have also been developed.^54^ However, all methods suffer from suboptimal accuracy and significant no-call rates^55^ and require as input personally identified information such as patient name and address, which makes them impractical in de-identified RWD settings. To address these shortcomings and take advantage of the molecular data present in clinico-genomic RWD, we previously developed an R/E imputation method that leverages genetic ancestry inferred from tissue sequence data and was reported to be of significantly higher accuracy than methods such as BISG^13^ This inclusion allows for larger sample sizes in the analysis of I/E criteria and yields more reliable data, while also protecting against the biases of R/E missingness. In applying race imputation methods, we followed established ethical imputation recommendations,^56^ auditing input data for bias, scrutinizing methodological choices to prevent bias introduction, and rigorously assessing the imputed data’s accuracy. Our adherence to these guidelines highlights our commitment to responsible race imputation use in promoting healthcare equity.^57^

An important consideration in applying this methodology is defining the disease under study. The FDA’s draft guideline on diversity plans suggests that enrollment goals should reflect the epidemiology of the targeted disease.^5,17^ In our example, the question arises: Is the disease being treated prostate adenocarcinoma, or specifically TMPRSS2:ERG+ prostate cancer? It has been suggested that early-onset metastatic and clinically advanced prostate cancer, characterized by a higher incidence of TMPRSS2:ERG fusions, is a distinct clinical and molecular entity.^58^ Racial disparities in the distribution of TMPRSS2:ERG fusions are well documented, with Black patients less likely to have these alterations compared to White patients.^59^ The causes of this difference are unclear, potentially due to genetic susceptibility, hormone levels, lifestyle factors, and healthcare access.^47^ The complexity of defining disease within cancer subtypes presents a challenge, particularly when incidence databases offer limited subtype-specific epidemiology.^60^ Given this complexity, we face a choice: set enrollments based on the distribution after applying I/E criteria and adjusting by factors derived from *WDF*, or make an additional adjustment to address biases introduced by selecting for the biomarker. While we did not make this additional adjustment in our example, it is up to clinical investigators and sponsors to decide, based on their specific aims and the nature of the therapy being tested. Adjusting R/E targets to compensate for disparities introduced by subtyping requires a delicate balance between scientific accuracy and equitable representation, highlighting the importance of precision in trial design.

Another point of consideration is when applying I/E criteria based on algorithmic scores that could be biased by R/E. For example, tumor mutational burden (TMB), a commonly used biomarker to predict the effectiveness of immunotherapy, is inflated in groups other than NH-White when determined from tumor-only sequencing.^61^ This artifact can be eliminated by matched tumor-normal analysis^61,62^ or empirically derived adjustment factors.^63^ Another example is the colorectal cancer consensus molecular subtypes (CMS) classifiers,^64^ which were developed mostly from data from White patients and may experience R/E biases and increased no-call rates in some groups.^65^ Care must be taken to assess whether these scores/classifiers are biased due to disparities in training data and the potential impact of such biases on R/E distributions.^16,66^

In our methodology, we introduce a scaling factor (α) to decide the extent of adjustment to the final R/E distribution based on the identified disparity between expected incidence data and the RWD cohort. Ideally, this factor is initially set to 1, to adjust according to the discovered disparities in our RWD. However, investigators have the flexibility to modify its impact based on ethical, statistical, and feasibility considerations. The key is balancing these adjustments to ensure ethical fairness and scientific validity in the recruitment process, which might involve iterative calculations and adjustments based on feedback from stakeholders and ethical guidelines.

Limitations of our study include our inability to impute R/E categories for underrepresented groups in our RWD such as American Indian or Alaska Native, and Native Hawaiian or Other Pacific Islander. Patients in these groups may receive no-call/”complex” calls or be misclassified into broader categories like Hispanic/Latino or Asian. Our R/E imputation method might require revalidation or retraining for use in other RWD sources or for updated federal R/E standards,^27^ and drift can occur over time as the US population’s admixture changes.^57^ Additionally, this imputation method is not directly applicable outside the US, where racial and ethnic categories differ and may encompass different genetic ancestries. Other limitations arise from averaging expected R/E distributions at the US state level, which may overlook potentially significant differences in cancer incidence at the county level due to environmental factors. This averaging is necessary to cope with the censoring that occurs in many counties for minority groups when patient counts are very low. Moreover, in de-identified data, we lack county-level patient addresses for similar privacy reasons. However, our intent is not to underscore such disparities but to match the RWD area of service as closely as possible to obtain accurate expectations. Additionally, our approach aims to define R/E enrollment targets based on the disease incidence, the disparities leading to the RWD, and the distortions in the R/E distribution introduced by using biomarkers. However, this does not guarantee that the final numbers of patients enrolled from different groups will allow for sufficiently powered subgroup analyses.^17^ If such analyses are desired, it may be necessary to supplement minority groups based on power calculations.

Setting R/E enrollment targets is a crucial aspect of a diversity plan, but it is only one part. A comprehensive strategy should also outline how to achieve these targets, such as by minimizing participation barriers for underrepresented minorities.^6,67–69^ This could involve opening trial sites in community practices, not just academic centers, educating patients and providers, and offering stipends, transportation, and telemedicine options to ease participation.^67^ Additionally, RWD can potentially assist in selecting sites where diverse patients are treated for specific cancers, and R/E imputation in RWD can provide a more complete and unbiased view of patient diversity at potential recruitment clinical sites.

## Conclusions

The FDORA legislation and FDA guidelines for diversity plans address the long-standing underrepresentation of minority groups in clinical trials, a crucial ethical concern in clinical research. Advocating for a data-driven approach, we emphasize utilizing real-world data (RWD), especially clinico-genomic databases in this endeavor. These databases, expanding significantly beyond resources such as the TCGA, offer unparalleled diversity and scale. By leveraging such insights, we can foster more inclusive clinical research and develop treatments that are safe and effective across all patient demographics.

## Data Availability

All data produced in the present work are contained in the manuscript.

## Acknowledgments

We thank Eric Schadt (Pathos AI), Matthew Conney, and Derrick Beech (Tempus AI) for reviewing the manuscript draft and providing valuable comments. We acknowledge Rafael Esleyer, Nick Riggan, and Arvind Prasad (Tempus AI) for their invaluable assistance in procuring the data needed for this work. Our gratitude extends to Frank Nothaft for his support with data access and to Joel Dudley, formerly of Tempus, for encouraging us to pursue this research. We also thank Vanessa Nepomuceno for her copy-editing of the manuscript.

## Authors’ contributions

FMDLV conceived the study, outlined the methods, and wrote the draft of the paper. FMDLV, YP and BR developed the statistical methodology. YP and BR obtained, processed data and performed statistical analysis. All authors edited and approved the manuscript.

## Ethical Approvals

All analyses were performed using safe-harbor de-identified data under the exemption Pro00042950 granted by Advarra, Inc. Institutional Review Board (IRB).

## Supplementary Materials

**Supplementary Figure 1.**
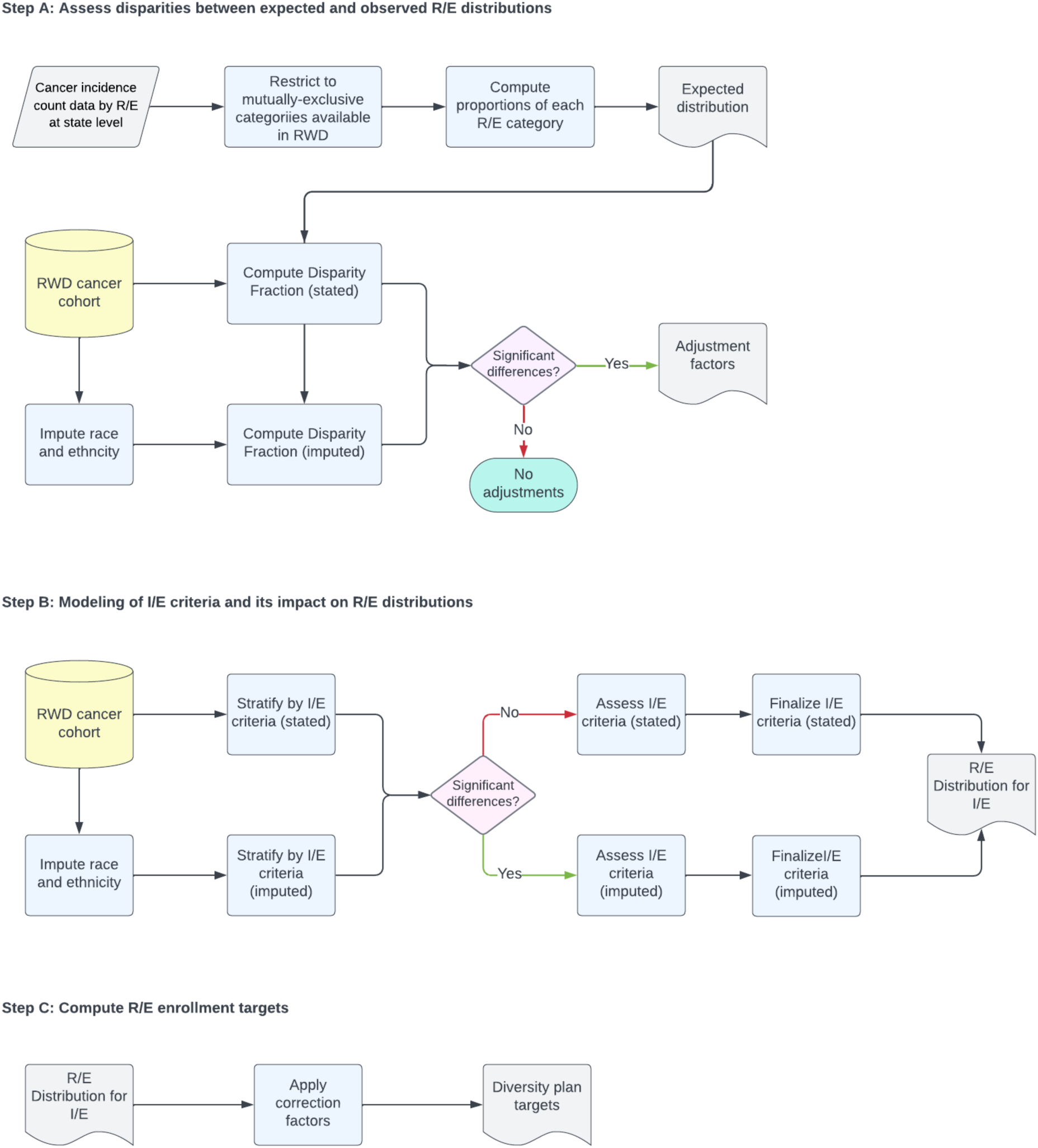
A workflow to establish R/E enrollment targets in oncology trials from clinico-genomic RWD.

**Supplementary Table 1.** Patient characteristics of prostate adenocarcinoma cohort by imputed race and ethnicity.

| Characteristic | NH Asian<br>N = 142 | NH Black<br>N = 772 | NH White<br>N = 2,893 | Hispanic/Latino<br>N = 389 |
| --- | --- | --- | --- | --- |
| Stated Race |  |  |  |  |
| White | 1 (1.4%) | 7 (1.8%) | 1,454 (98%) | 58 (51%) |
| American Indian or<br>Alaska Native | 0 (0%) | 0 (0%) | 1 (<0.1%) | 4 (3.5%) |
| Asian | 61 (88%) | 0 (0%) | 1 (<0.1%) | 0 (0%) |
| Black or African<br>American | 1 (1.4%) | 383 (97%) | 0 (0%) | 10 (8.8%) |
| Native Hawaiian or<br>Other Pacific Islander | 1 (1.4%) | 0 (0%) | 0 (0%) | 0 (0%) |
| Other Race | 5 (7.2%) | 4 (1.0%) | 26 (1.8%) | 42 (37%) |
| Race not stated | 0 (0%) | 1 (0.3%) | 2 (0.1%) | 0 (0%) |
| Unknown | 73 | 377 | 1,409 | 275 |
| Stated ethnicity |  |  |  |  |
| Not Hispanic or Latino | 47 (100%) | 191 (95%) | 903 (99%) | 24 (15%) |
| Hispanic or Latino | 0 (0%) | 11 (5.4%) | 12 (1.3%) | 135 (85%) |
| Unknown | 95 | 570 | 1,978 | 230 |
| Age at collection |  |  |  |  |
| Present | 68 (63, 75) | 64 (59, 69) | 68 (62, 74) | 64 (60, 70) |
| Unknown | 14 | 86 | 368 | 29 |
| Age at Dx |  |  |  |  |
| Present | 68 (63, 76) | 64 (59, 69) | 67 (61, 73) | 64 (59, 70) |
| Unknown | 29 | 142 | 572 | 62 |
| Max. total Gleason score |  |  |  |  |
| 6 | 1 (0.8%) | 10 (1.4%) | 21 (0.8%) | 2 (0.6%) |
| 7 | 23 (18%) | 135 (20%) | 463 (18%) | 69 (19%) |
| 8 | 34 (27%) | 147 (21%) | 505 (20%) | 70 (19%) |
| 9 | 54 (42%) | 338 (49%) | 1,312 (52%) | 189 (53%) |
| 10 | 16 (12%) | 60 (8.7%) | 226 (8.9%) | 29 (8.1%) |
| Unknown | 14 | 82 | 366 | 30 |

## Notes

### Competing Interest Statement

F.M.D.L., Y.P., and B.R. and are employees and have received stock options from Tempus AI, Inc.

### Funding Statement

This study was funded by Tempus AI, Inc.

### Author Declarations

The Institutional Review Board of Advarra, Inc., waived ethical approval for this work.

